# Improving the quality of venous blood sampling procedure (phlebotomy): avoiding tourniquet use

**DOI:** 10.1101/2020.04.05.20039560

**Authors:** Francisco Freitas, Mónica Alves

## Abstract

**Background:** Guidelines for venous blood sampling procedure (phlebotomy) discourage tourniquet use whenever possible. Here, we aimed to assess the Biomedical Scientists capability of not using the tourniquet in phlebotomy, which we hypothesized to be equal to 50% of the patients attended, and identifying the most frequent venipuncture site.

**Materials and Methods:** We selected and assigned two (BMS) with the same age (41 years) and experience (20 years) to record ten phlebotomy days, the first with prioritized and the latter with non-prioritized patients. In a simple record form, each acquired daily data for the number of attended patients, age and gender, the frequency of non-tourniquet usage and the punctured vein. To test our work hypothesis we used the two-tailed single sample t-test (p < 0.05). Differences between age-group means and non-tourniquet use means by each BMS were tested by two-tailed t-test for independent means (p < 0.05).

**Results:** In 10 phlebotomy days 683 patients were attended, with males representing 43,2% of the population. We found no statistically difference between age-group means. The combined capability of non-tourniquet use was 50,5%, which did not differ from our null hypothesis, but the individual group-means were statistically different, being 33% and 66.9% in the prioritized vs non-prioritized group. The medial cubital vein was the most prone to be punctured (77,7%).

**Conclusions:** We have shown that performing phlebotomies without tourniquet use is possible and desirable in at least half of the attended patients, though being more limited in specific group populations. Our results provide room for quality improvement in the laboratory pre-analytical phase.

**Key points summary:** We assessed the capability of Biomedical Scientists not using the tourniquet in real life blood sampling procedures for diagnostic purposes.

Blood was collected from at least half of the attended patients without tourniquet use.

Biomedical Scientists were able to prioritize the antecubital veins without tourniquet application (medial cubital vein the most prone to be punctured - 78% of attempts).

## 1. Introduction

Phlebotomy (venous blood sampling) is probably the most common invasive procedure performed in health-care, being a fundamental basis for surveillance, diagnosis and treatment of patients, and is part of the pre-analytical phase (before the sample is analysed in a laboratory). In Portugal, phlebotomy is a part of the routine day work mainly of nonmedical personnel like nurses and Biomedical Scientists (BMS), also known as Medical Laboratory Technicians. In an ageing world, this procedure is becoming more frequent and sometimes more difficult due to patient multiple morbidities and/or disabilities.

Several guidelines or recommendations are available to standardize venous blood sampling practice, like the Joint EFLM-COLABIOCLI [1] or the CLSI GP41-A7 standard [2], with both having a detailed step-by-step approach.

Here we point out to the tourniquet application step, which we usually use less than a minute to constrict venous circulation, gaining better vein location and access. Both of the above guidelines discourage its use whenever possible, due to the effects on haemoconcentration, which can lead to spurious results [3-6] or possible hemolysis due to prolonged venous stasis [7,8], thus impacting on patient safety and results turnaround time.

Lima-Oliveira and co-workers have published on tourniquet application time and demonstrated that the CLSI H3-A6 standard [9] induces increased application time [10,11], which led to a modification proposal on the procedure by applying the tourniquet on step IX rather than on step VI [4]. Still, the actual GP41-A7 standard recommend putting gloves on after applying tourniquet. It also declares that the phlebotomist may not be able to prioritize the antecubital veins without tourniquet application [2]. In our laboratory, BMS are sensitive to tourniquet application time and proceed to early release after needle insertion, but only a few are prone to let the tourniquet out of process whenever possible. In light of this, and since there is a gap of data in the literature, the aim of the present study was to assess the Biomedical Scientists capability of not using the tourniquet in real life blood sampling procedures for diagnostic purposes. We hypothesised that our capability of non-tourniquet use equals 50% of attended patients. We also aim to identify the most frequent venipuncture site without tourniquet use.

## 2. Materials and Methods

### 2.1 Subjects and Materials

Our laboratory has five boxes for blood sampling procedures, three for non-prioritized and two for prioritized patients. In the first group, we usually attend general outpatients with a first medical consultation or in follow-up, for an elective surgery or under oral anticoagulants. In the second group, the Portuguese law stands that public hospitals should gave priority to outpatients with physical (in wheel chair / litter/ other) or mental disabilities, pregnant women, and people accompanied by infants. Our laboratory also prioritize patients with type I diabetes, with prescribed exams made in an external laboratory or another following exam in the hospital, and children (6-12 years). Other outpatients are attended only after these.

To standardize the evaluation of tourniquet application and reduce bias between patient-groups, we selected and assigned two biomedical scientists (BMS) with the same age (41 years) and experience (20 years), one for each group.

No informed consent or ethical approval was required for this study, as no specific patient information is presented.

### 2.2 Methods

In a two-day practical coaching period, BMS were instructed to avoid tourniquet use whenever possible, by asking every patient to make a fist first (without pumping), in order to assess the presence of prominent veins (prioritizing antecubital), and the possibility of a direct phlebotomy. When blood flow starts, the fist shall then be open. If not possible or confident enough to perform, phlebotomy should be made using the tourniquet. In general, our blood sampling procedure follows the steps of EFLM-COLABIOCLI recommendations [1].

The study was conducted between April 2^nd^ and May 10^th^ 2019, a period in which both BMS recorded ten phlebotomy days. In a simple record form, each acquired daily data for the number of attended patients, age and gender, the frequency of non-tourniquet usage and the respectively punctured vein.

### 2.3 Statistical analysis

To test our work hypothesis, we used relative and combined frequencies of the attended patients without tourniquet use, and tested the difference with the two-tailed single sample t-test (p < 0.05). The Kolmogorov-Smirnov test was used to assess the normality of distribution of investigated variables (age and non-tourniquet use), which were considered to be normally distributed. Differences between age-group means and non-tourniquet use means by each BMS were tested by two-tailed t-test for independent means (p < 0.05). To present data on the punctured vein without tourniquet use we used absolute count and relative frequencies. Statistical analysis was done using SPSS version 20 software (IBM Corporation, Armonk, NY, USA).

## 3. Results

The main results of the present study are summarized in Table 1. In 10 phlebotomy days both BMS attended 683 patients (plus 23 in non-priority group), with males representing 43,2% of the total population. We found no statistically difference between age-group means (p = 0.334). Considering the non-tourniquet use, we verified that the individual means were statistically different (p = <0.001), with BMS 1 having all of ten 10 days above 53%, while BMS 2 had only ∼42% in two days. The combined capability of non-tourniquet use was shown to be 50,5% (345 out of 683 attended patients), which did not differ from our null hypothesis (p = 0.909). None of the BMS reported a failed phlebotomy when not using the tourniquet.

**Table 1.** Population study demographics and non-tourniquet-use (NTU) in attended-groups by each BMS.

| Day | BMS 1 |  |  |  |  | BMS 2 |  |  |  |  |
| --- | --- | --- | --- | --- | --- | --- | --- | --- | --- | --- |
|  | Non-priority group |  |  |  |  | Priority group |  |  |  |  |
|  | TP | Male | Age (years) | NTU<br>N | NTU<br>% | TP | Male | Age (years) | NTU<br>N | NTU<br>% |
| 1 | 35 | 18/35 | 58 (18-85) | 20 | 57,1 | 27 | 10/27 | 57 (11-80) | 7 | 26 |
| 2 | 36 | 15/36 | 55 (8-93) | 20 | 55,5 | 37 | 19/37 | 57 (18-91) | 7 | 19 |
| 3 | 34 | 14/34 | 61 (8-92) | 23 | 67,6 | 34 | 16/34 | 55 (20-95) | 8 | 23,6 |
| 4 | 34 | 14/34 | 54 (16-83) | 21 | 61,7 | 28 | 10/28 | 60 (21-88) | 9 | 32,1 |
| 5 | 34 | 16/34 | 54 (13-86) | 27 | 79,4 | 36 | 17/36 | 58 (17-85) | 13 | 36,1 |
| 6 | 30 | 13/30 | 59 (16-91) | 16 | 53,3 | 27 | 16/27 | 61 (21-84) | 10 | 37 |
| 7 | 28 | 13/28 | 57 (34-81) | 15 | 53,6 | 24 | 6/24 | 59 (29-85) | 10 | 41,6 |
| 8 | 43 | 21/43 | 59 (27-83) | 38 | 88,3 | 34 | 14/34 | 60 (12-93) | 13 | 38,2 |
| 9 | 39 | 18/39 | 63 (33-88) | 28 | 71,8 | 37 | 12/37 | 63 (21-92) | 13 | 35,1 |
| 10 | 40 | 17/40 | 60 (20-86) | 28 | 70 | 46 | 16/46 | 63 (11-96) | 19 | 41,3 |
| Total | 353 | 159/353 | 58 (8-93) | 236 | 66,9 | 330 | 136/330 | 59 (11-96) | 109 | 33 |

TP – Total patients attended. NTU N – Non-Tourniquet-Use absolute count.

As stated in Table 2, the medial cubital vein was the most prone (77,7%) to be punctured when tourniquet was not used, followed by the basilica and cephalic veins respectively, with both BMS presenting the same pattern.

**Table 2.**
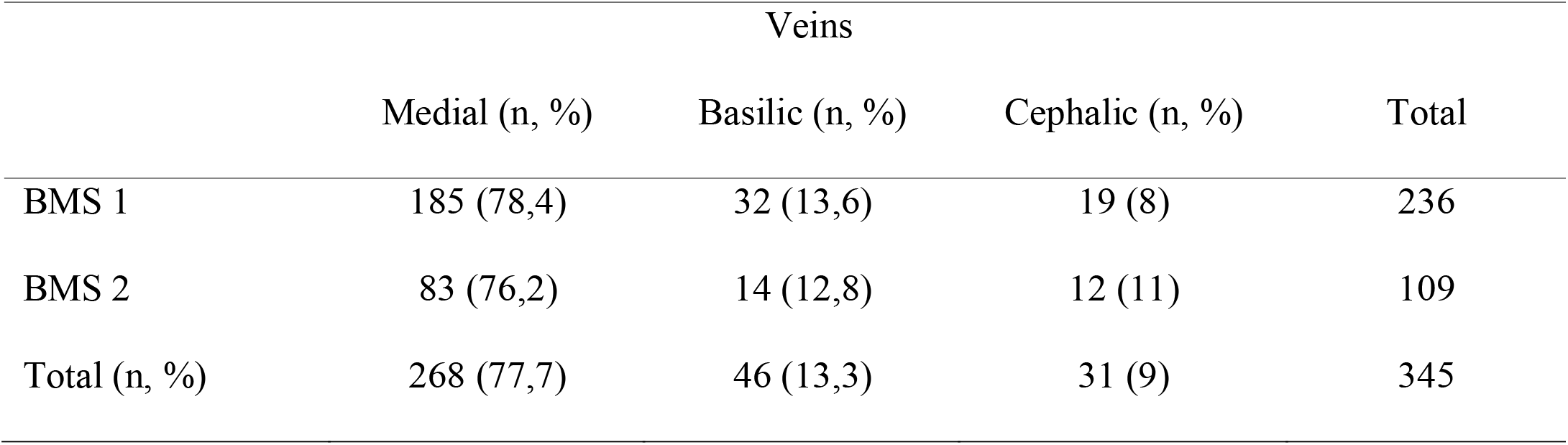
Punctured veins by each BMS without tourniquet use.

## 4. Discussion

Tourniquet use in blood sampling procedures has been a source for several studies, mainly directed to its potential to produce spurious analytical results and impact on patient safety. Current evidence suggests that prolonged tourniquet application time (> 1 min) can lead to results exceeding the current analytical quality specifications for desirable bias, either in biochemical (total protein, albumin, potassium and calcium) or heamathological testing (WBC, RBC, hemoglobin, hematocrit, lymphocytes and monocytes) [5,6,12,13]. Interestingly, platelet function assessed by multiple electrode aggregometry [3], and erythrocyte deformability and aggregation also seem to be significantly affected [14,15]. Higher rates of hemolyzed samples are also associated with tourniquet application times greater than one minute [7,8], leading to rejection and new sample request, impacting on tests turnaround times.

In face of the raised issues, a new device to locate veins by transillumination technology has been tested, and proved to be a useful tool by eliminating the venous stasis and improving the quality of phlebotomy procedures, especially critical in patients with difficult or small veins, such as children and old people [5,16]. Despite the advantages, this technology has not paved its way into the health-care setting, which is also our case scenario.

To our knowledge, this is the first study reporting on the real capability of non-tourniquet use when performing a phlebotomy, which we have shown to be 50,5% of attended patients, confirming our working hypothesis. This value will set the benchmark for future studies on this problematic. Since our laboratory has three boxes for non-prioritized and two for prioritized patients, with appropriate educational interventions we can project a potentially non-tourniquet phlebotomy ratio well >50%. The medial cubital vein was paramount to our results (Table 2), because it’s the most prominent and easy to puncture, usually doesn’t roll under the skin, and is also a good choice to prevent nerve injury and an hemolyzed sample [1, 17-19]. Currently there is no study with which we can compare our results, but as stated by Lima-Oliveira et al, about 78% of phlebotomies in outpatients are performed in the medial cubital vein when using a tourniquet [20], and we have achieved similar results without its use.

Our results are in clear contradiction with the GP41-A7 standard statement, declaring that the phlebotomist may not be able to prioritize the antecubital veins without tourniquet application [2]. Adding to this, neither BMS had a failed phlebotomy, which demonstrates good selection and confidence to perform.

We have also confirmed that performing phlebotomy in prioritized patients limits our capability of drawing blood without tourniquet use, despite no difference found between age-group means. This clearly shows that we stand before true-different group-populations.

Three limitations need to be acknowledged regarding the present study. The first is that each phlebotomist as its own technique and confidence to perform, and we cannot measure its effect on the results. The second limitation is the number of phlebotomy days. As stated in Table 1, BMS 2 had better and maintained values of non-tourniquet use since day 5. The third one is that we have not tested the reverse group-BMS pair to verify that the results were still concordant.

## 5. Conclusions

This is the first study reporting on the real capability of non-tourniquet use when performing a phlebotomy, which we have shown to be possible and desirable in at least half of the attended patients, though being more limited in specific group populations. We also demonstrate that BMS were able to prioritize the antecubital veins without tourniquet application, with the medial cubital vein the most prone to be punctured (78% of attempts). Our results clearly provide some room for continuous quality improvement in the laboratory pre-analytical phase, which is one of the primary sources of errors in the total testing process.

## Data Availability

All data is readilly available.

## Declarations

### Authors Contributions

FF and MA both declare:

1. Made a substantial contribution to the concept or design of the work; or acquisition, analysis or interpretation of data;
2. Drafted the article or revised it critically for important intellectual content;
3. Approved the version to be published;
4. Participated sufficiently in the work to take public responsibility for appropriate portions of the content.
5. The manuscript is not under review of any other journal, and that it has not been published complete or partially in any other jornal, though a previous version has already a DOI in medRxiv (doi.org/10.1101/2020.04.05.20039560)

## Ethics Approval

Not applicable

## Acknowledgements

None to declare

## References

1. Simundic AM, Bölenius K, Cadamuro J, Church S, Cornes MP, van Dongen-Lases EC, et al. Joint EFLM-COLABIOCLI Recommendation for venous blood sampling. Working Group for Preanalytical Phase (WG-PRE), of the European Federation of Clinical Chemistry and Laboratory Medicine (EFLM) and Latin American Working Group for Preanalytical Phase (WG-PRE-LATAM) of the Latin America Confederation of Clinical Biochemistry (COLABIOCLI). Clin Chem Lab Med 2018;56(12):2015–38.

2. Clinical and Laboratory Standards Institute. GP41: Procedures for the Collection of Diagnostic Blood Specimens by Venipuncture; Approved Standard—Seventh Edition. CLSI document GP41-A7. Wayne, PA: Clinical and Laboratory Standards Institute, 2017.

3. Lima-Oliveira G, Lippi G, Salvagno GL, Gaino S, Poli G, Gelati M, et al. Venous stasis and whole blood platelet aggregometry: a question of data reliability and patient safety. Blood Coagul Fibrinolysis 2015;26(6):665–8.

4. Lima-Oliveira G, Lippi G, Salvagno GL, Montagnana M, Picheth G, Guidi GC. The effective reduction of tourniquet application time after minor modification of the CLSI H03-A6 blood collection procedure. Biochem Med 2013;23(3):308–15.

5. Lima-Oliveira G, Lippi G, Salvagno GL, Montagnana M, Manguera CL, Sumita NM. New ways to deal with known preanalytical issues: use of transilluminator instead of tourniquet for easing vein access and eliminating stasis on clinical biochemistry. Biochem Med 2011;21:152–9.

6. Lippi G, Salvagno GL, Montagnana M, Brocco G, Guidi GC. Influence of short-term venous stasis on clinical chemistry testing. Clin Chem Lab Med 2005;43(8):869–875.

7. Phelan MP, Reineks EZ, Schold JD, Hustey FM, Chamberlin J, Procop GW. Preanalytic Factors Associated With Hemolysis in Emergency Department Blood Samples. Arch Pathol Lab Med 2018;142(2):229–235.

8. McCaughey EJ, Vecellio E, Lake R, Li L, Burnett L, Chesher D, et al. Key factors influencing the incidence of hemolysis: A critical appraisal of current evidence. Crit Rev Clin Lab Sci 2017;54(1):59–72.

9. Clinical and Laboratory Standards Institute. Procedures for the Collection of Diagnostic Blood Specimens by Venipuncture CLSI H3-A6 document. 6th ed. Wayne, PA: Clinical Laboratory Standards Institute, 2007.

10. Lima-Oliveira G, Lippi G, Salvagno GL, Montagnana M, Picheth G, Guidi GC. Impact of the phlebotomy training based on CLSI/NCCLS H03-A6 - procedures for the collection of diagnostic blood specimens by venipuncture. Biochem Med 2012;22(3):342–51.

11. Lima-Oliveira G, Guidi GC, Salvagno GL, Montagnana M, Rego FGM, Lippi G, et al. Is phlebotomy part of the dark side in the clinical laboratory struggle for quality? Lab Med 2012;43:172–6.

12. Lippi G, Salvagno GL, Montagnana M, Franchini M, Guidi GC. Venous stasis and routine hematologic testing. Clin Lab Haematol 2006;28:332–7.

13. Lippi G, Salvagno GL, Solero GP, Guidi GC. The influence of the tourniquet time on hematological testing for antidoping purposes. Int J Sports Med 2006;27(5):359–62.

14. Cengiz M, Ulker P, Meiselman HJ, Baskurt OK. Influence of tourniquet application on venous blood sampling for serum chemistry, hematological parameters, leukocyte activation and erythrocyte mechanical properties. Clin Chem Lab Med 2009;47(6):769–76.

15. Connes P, Uyuklu M, Tripette J, Boucher JH, Beltan E, Chalabi T, et al. Sampling time after tourniquet removal affects erythrocyte deformability and aggregation measurements. Clin Hemorheol Microcirc 2009;41(1):9–15.

16. Lima-Oliveira G, Lippi G, Salvagno GL, Montagnana M, Scartezini M, Guidi GC, et al. Transillumination: a new tool to eliminate the impact of venous stasis during the procedure for the collection of diagnostic blood specimens for routine haematological testing. Int J Lab Hematol 2011;33(5):457–62.

17. Ohnishi H. How to Prevent Phlebotomy-Related Nerve Injury. Rinsho Byori 2007;55(3):251– 6.

18. Mukai K, Nakajima Y, Nakano T, Okuhira M, Kasashima A, Hayashi R, et al. Safety of Venipuncture Sites at the Cubital Fossa as Assessed by Ultrasonography. J Patient Saf 2020;16(1):98–105.

19. Lippi G, Avanzini P, Cervellin G. Blood collection from intravenous lines: is one drawing site better than others? Lab Med 2014;45(2):172–5.

20. Lima-Oliveira G, Lippi G, Salvagno GL, Picheth G, Guidi GC. Laboratory Diagnostics and Quality of Blood Collection. J Med Biochem 2015;34(3): 288–94.

